# Comparison of blood and urine concentrations of equol by LC‒MS/MS method and factors associated with equol production in 466 Japanese men and women

**DOI:** 10.1101/2023.07.10.23292459

**Authors:** Remi Yoshikata, Khin Zay Yar Myint, Junichi Taguchi

## Abstract

Equol is produced from daidzein by the action of gut bacteria on soy isoflavones. However, not all people can produce equol, and metabolism differs even among the producers. We aimed to examine the equol producer status in both men and women, and investigate the relationships among the serum and urinary isoflavones as well as to other biomedical parameters. In this study, we measured the equol and daidzein concentrations from the blood and urine of 292 men and 174 women aged between 22 and 88 years by liquid chromatography/tandem mass spectrometry (LC/MS/MS).We then analysed the cut-off value for equol producers in both sexes, the relationship of serum and urinary equol concentrations, and other parameters, such as sex, age, endocrine function, glucose metabolism, lipid metabolism, and renal function with regards to equol-producing ability, among the different age groups. Equol producers were defined as those whose log ratio of urinary equol and daidzein concentration or log (equol/daidzein) was -1.42 or higher. Among 466 participants, 195 were equol producers (42%). The proportion of equol producers was larger in women. The cut-off value for equol producers was consistent in both sexes. Positive relationships were noted between serum and urinary equol levels in equol producers of both sexes; however, such an association was not detected in nonproducers. PSA levels in men were significantly lower in equol producers (0.8 v.s. 1.0 ng/ml, p=0.004), especially in those in their 40 s (0.82 vs. 1.13 ng/ml, p<0.001) and 60 s (0.64 vs. 1.02 ng/ml, p<0.001).

## Introduction

Dietary isoflavones are metabolized in the lower part of the small and large intestine into three main ingredients of soy isoflavones: daidzein, genistein and glycitein. There, the end-product or active metabolite of daidzein called ‘equol’ could be produced only in certain people who have functional equol-producing bacteria. Equol then enters the blood stream after being absorbed from the intestinal wall and is distributed to the target organs. Similar to other dietary end-products, it is chemically modified in the liver and mainly excreted in the urine, with only a small amount excreted into the stool. It can be detected in the blood 8 hours after ingestion of isoflavones and reaches a peak concentration after 12 to 24 hours. After 72 hours, its blood level becomes negligible, as it is rarely reserved in the body [1].

Equol exists as enantiomers, R-equol and S-equol. However, S-equol is the only product that can be identified in the blood and urine of humans and animals [2–3]. It has the highest potential for inducing health benefits among soy isoflavones, as it possesses estrogenic and antiestrogenic actions as well as antiandrogenic and antioxidant actions [4–6]. It is associated with the relief of menopausal symptoms and a reduced risk of related conditions, including osteoporosis [7–11], in women. For men, it was found to be associated with a reduced risk of prostate cancers [12,13]. In addition, it was reported to have positive and antiaging effects on skin structures in both men and women [14, 15].

However, not everyone has the ability to produce equol [2]. The ability to produce equol depends on age, gender, genetics, dietary contents, and other factors [16–18]. Therefore, health benefits are not observed in some people even if they consume soy isoflavones. Even in those with equol-producing ability, individual and diurnal variations and the use of antibiotics greatly influence the desirable level of equol in the body [19–21]. For those reasons, there have been attempts to induce equol actions through supplements containing equol or with probiotics such as Lactobacilli and Bifidobacteria [22].

We hypothesized that there were differences between men and women with regards to equol producing ability, as well as its relationship with other biomarkers inside the body. In this study, we aimed to examine cut-off value of equol producers, the relationship of blood and urinary equol, as well as their relationships with other parameters, such as sex, age, endocrine function, glucose metabolism, lipid metabolism, and renal function, in both sexes among the different age groups.

## Materials and Methods

### Participants

This cross-sectional study was conducted among 466 healthy men and women (292 men and 174 women) aged between 22 and 88 (mean age 55) years who were receiving annual health screening at the Himedic Kyoto University Hospital from June 2016 to December 2017.

### Ethical considerations

We included the data of the participants from all the men and women within the study period who provided general consent (Supplementary data file 1) for the use of their health screening data for secondary purposes. The study was approved by the Institutional Review Board of The University of Tokyo (Supplementary data file 2).

### Measurements

Equol and daidzein concentrations in the serum and urine were determined by the liquid chromatography/tandem mass spectrometry (LC/MS/MS) method, measured by LSI Medience Corporation (Tokyo, Japan) using Multiple Reaction Monitoring Triple quadrupole mass spectrometry (LCMS-8050, Shimadzu, Japan). The limits of detection (LODs) were 1 ng/mL for serum equol and daidzein, 10 ng/mL for urine equol, and 20.0 ng/mL for urine daidzein. Liquid chromatography‒mass spectrometry (LC‒MS) is an indispensable tool for quantitative and qualitative analysis in a wide range of fields, from pharmaceuticals and food science to environmental analysis. It has been used extensively in quantitative measurements of isoflavones in several studies [23–27]. Based on a previous epidemiological study in 4,412 Japanese women [28], equol producers were defined as those whose log ratio of urinary equol and daidzein concentration or log (equol/daidzein) was -1.42 or higher.

### Metabolic parameters

Blood samples were obtained after an overnight fast to determine the fasting blood glucose level(FBG), Hemoglobin A1c (HbA1c), Glycated albumin, 1.5-Anhydro-D-glucitol, Insulin, C-peptide, Homeostatic Model Assessment for Insulin Resistance (HOMA-IR), total cholesterol, low density lipoprotein cholesterol (LDL-C), triglycerides(TG), high density lipoprotein cholesterol (HDL-C), uric acid(UA), urinary creatinine(UCRE), high sensitivity C-reactive protein (hs-CRP), Estradiol(E2), Luteinizing hormone(LH), Follicle stimulating hormone (FSH), Thyroid stimulating hormone (TSH), free-Triiodothyronine(f-T3), free- Thyroxine(f-T4).

### Statistical analysis

We first calculated the effected size for equol producer proportions by gender using Setchell’s study on 23 men and 18 women, with equol producer proportions of 65% for men and 28% for women. Using that effect size (w=0.162069), with the assumption of a two-sided 5% significance level and power of 80%, and calculated that a minimum of 415 subjects in total was necessary for the comparison of equol producers in gender by Chi-squared test. Considering 10% opt-out rate, we included data from 466 subjects. Serum and urinary S-equol and daidzein concentrations were expressed as micrograms/liter and when analysed were expressed as medians and interquartile ranges. The ratio of urinary equol to daidzein was calculated, transformed, and expressed as log10. We assessed the distributions of log10- transformed urinary equol:daidzein concentrations in histograms and used mixed models to examine any difference between the sexes to determine the equol producer status. According to the results, subjects with a ratio above −1.42 on this scale were classified as equol producers. We calculated the creatinine-corrected values of isoflavones to examine the associations between serum and urinary equol concentrations by linear regression analysis. The distribution of normality of the parameters was assessed with the Kolmogorov–Smirnov test, box plots, and histograms. We then compared the differences in parameters with regard to equol-producing ability using the Mann‒Whitney U test. The proportions of abnormal values were compared using the chi-squared test. All continuous variables are expressed as medians and interquartile ranges, and categorical variables are expressed as numbers and percentages. The calculations and figure generation, except for the generation of the mixed model histograms, were performed using Microsoft Excel (Microsoft Corporation, 2019). The sample size calculations, and mixed model histograms were generated using R software (R 4.3.0, R Core Team, 2023). All tests were two-sided, and statistical significance was set to p < 0.05.

## Results

### Evaluation of equol producer status

We applied the same finite fixed model as the previous study to determine the optimal cut-off value to distinguish between equol producers and nonproducers. The log10-transformed ratios of urinary equol to daidzein concentrations were plotted as shown in **Figure 1** across all participants (n=466), as well as in male (n=292) and female (N=174) participants separately. In all distributions, we observed that the cut-off values were approximately -1.4, without significant distinction between the male and female participants. This was consistent with our definition of equol producers for both sexes, i.e., at the value of -1.42. Therefore, our definition of equol producers as those having a log 10-transformed ratio of urinary equol to daidzein of - 1.42 or higher was relevant in our study population. Based on this definition, among 466 participants, 195 were equol producers (42%). The proportion of equol producers in women was 47%, whereas that in men was 39%.

**Figure 1.**
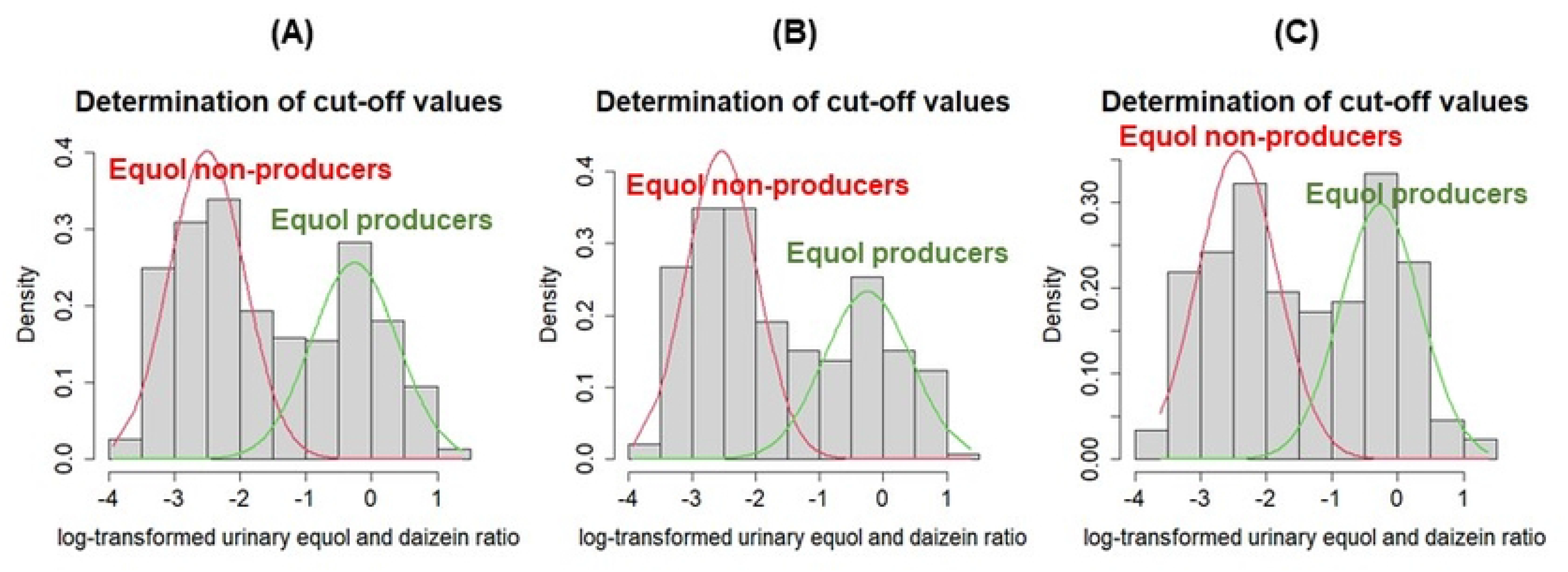
Assessing the cut-off values for Log10-transformed urinary equol: daidzein ratios. Determination of cut-off values in (A) all participants (N = 466), (B) males (N = 292), and (C) females (N = 174) using mixed model analysis. In all distributions, the cut-off values were approximately -1.4, without significant distinction between the male and female participants, i.e., at the value of -1.42.

### Comparisons of parameters between male and female participants

**Table 1** shows that female participants had higher equol concentrations in both serum and urine, glycated albumin, HDL-C, E2, LH, and FSH, which is consistent with other studies. Male participants had higher fasting blood glucose, 1.5-Anhydro-D-glucitol, C-peptide, HOMA-IR, TG, UA, UCRE, f-T3 and f-T4.

**Table 1:** Comparisons of parameters between male and female participants.

|  | Male (N=292) |  | Female (N=174) |  | p value |
| --- | --- | --- | --- | --- | --- |
| Equol producers, n (%) | 114 | (39%) | 81 | (47%) | 0.136<sup>a</sup> |
| Age quartiles (years) | (22 – 86) |  | (28 – 88) |  | 0.377<sup>a</sup> |
| 20s, n (%) | 2 | (1%) | 2 | (1%) |  |
| 30s, n (%) | 31 | (11%) | 19 | (11%) |  |
| 40s, n (%) | 67 | (23%) | 40 | (23%) |  |
| 50s, n (%) | 84 | (29%) | 61 | (35%) |  |
| 60s, n (%) | 82 | (28%) | 31 | (18%) |  |
| 70s, n (%) | 23 | (8%) | 15 | (9%) |  |
| 80s, n (%) | 3 | (1%) | 6 | (3%) |  |
| Serum equol (µg/gCr) | 0.6 | (0.34, 3.37) | 1.03 | (0.61, 7.23) | **<0.001** |
| Serum daidzein (µg/gCr) | 11.74 | (3.96, 35.35) | 17.85 | (7.16, 44.36) | **0.004** |
| Urinary equol (µg/gCr) | 7.52 | (3.7, 209) | 14.11 | (6.8, 600.8) | **<0.001** |
| Urinary daidzein (µg/gCr) | 1490.57 | (472.11, 4275.9) | 1876.56 | (761, 5997) | 0.07 |
| log (Equol/Daidzein) | -1.99 | (-2.74, 4275.9) | -1.59 | (-2.49, -0.3) | 0.121 |
| E2 | 0 | (0, 0) | 0.25 | (0.25, 66.55) | **<0.001** |
| LH | 0 | (0, 5.13) | 22.55 | (7.1, 32.68) | **<0.001** |
| FSH | 0 | (0, 5.78) | 47.65 | (7.48, 70.23) | **<0.001** |
| CRP | 0.05 | (0.05, 0.1) | 0.05 | (0.05, 0.1) | 0.167 |
| TSH | 1.65 | (1.1, 2.51) | 1.83 | (1.21, 2.63) | 0.106 |
| FT4 | 1.28 | (1.18, 1.4) | 1.21 | (1.13, 1.32) | **<0.001** |
| FT3 | 3.14 | (2.88, 3.41) | 2.8 | (2.63, 3.06) | **<0.001** |
| Glucose | 94 | (88, 103) | 90 | (85.3, 95.75) | **<0.001** |
| HbA1C | 5.60% | (0.05, 0.06) | 5.70% | (0.05, 0.06) | 0.814 |
| Glycated albumin | 0.13 | (0.13, 0.14) | 0.14 | (0.13, 0.15) | **<0.001** |
| 1.5-Anhydro-D-glucitol | 18.7 | (13.48, 22.53) | 15.95 | (12.8, 19.48) | **<0.001** |
| Insulin | 5 | (3.38, 7.6) | 4 | (2.7, 5.3) | 0.423 |
| C-peptide | 1.62 | (1.16, 2.08) | 1.16 | (0.91, 1.59) | **<0.001** |
| HOMA-IR | 1.17 | (0.76, 1.85) | 0.91 | (0.6, 1.32) | **<0.001** |
| Insulin resistance, n (%) | 40 | (14%) | 14 | (8%) | 0.093<sup>a</sup> |
| Total cholesterol | 197.5 | (175.8, 226) | 202 | (181, 223) | 0.289 |
| LDL | 115 | (99, 138) | 116.5 | (97, 137) | 0.878 |
| TG | 114.5 | (74, 177.25) | 69.5 | (53.3, 106.5) | **<0.001** |
| HDL | 55 | (47, 65) | 69 | (55.3, 78) | **<0.001** |
| UA | 6.15 | (5.4, 6.9) | 4.65 | (3.9, 5.38) | **<0.001** |
| UCRE | 122 | (80.75, 173) | 72 | (50.3, 111.5) | **<0.001** |

Continuous variables are expressed as medians (interquartile ranges) and were compared by the Mann–Whitney U test except for \*: chi-squared test; statistically significant p values are bold.

### Relationship between serum and urine equol levels

Figure 2 showed strong positive relationships between the serum and urine equol levels in both male (r= 0.75, R^2^ = 0.56, p<0.001) and female equol producers (r=0.63, R^2^ = 0.39, p<0.001). However, such an observation was not observed in equol nonproducers in either sex (r=0.24, R^2^ = 0.0576, p<0.01, and r=0.03, R^2^ = 0.0008, p=0.8, respectively). Equol nonproducers tended to have greater variances of urinary equol with reference to serum equol concentration, or they had almost no relationship between these two parameters. The relationship between serum and urinary equol concentrations was weaker in female equol producers, as their regression slope was lenient than that of male equol producers, although there were some individual differences.

**Fig 2.**
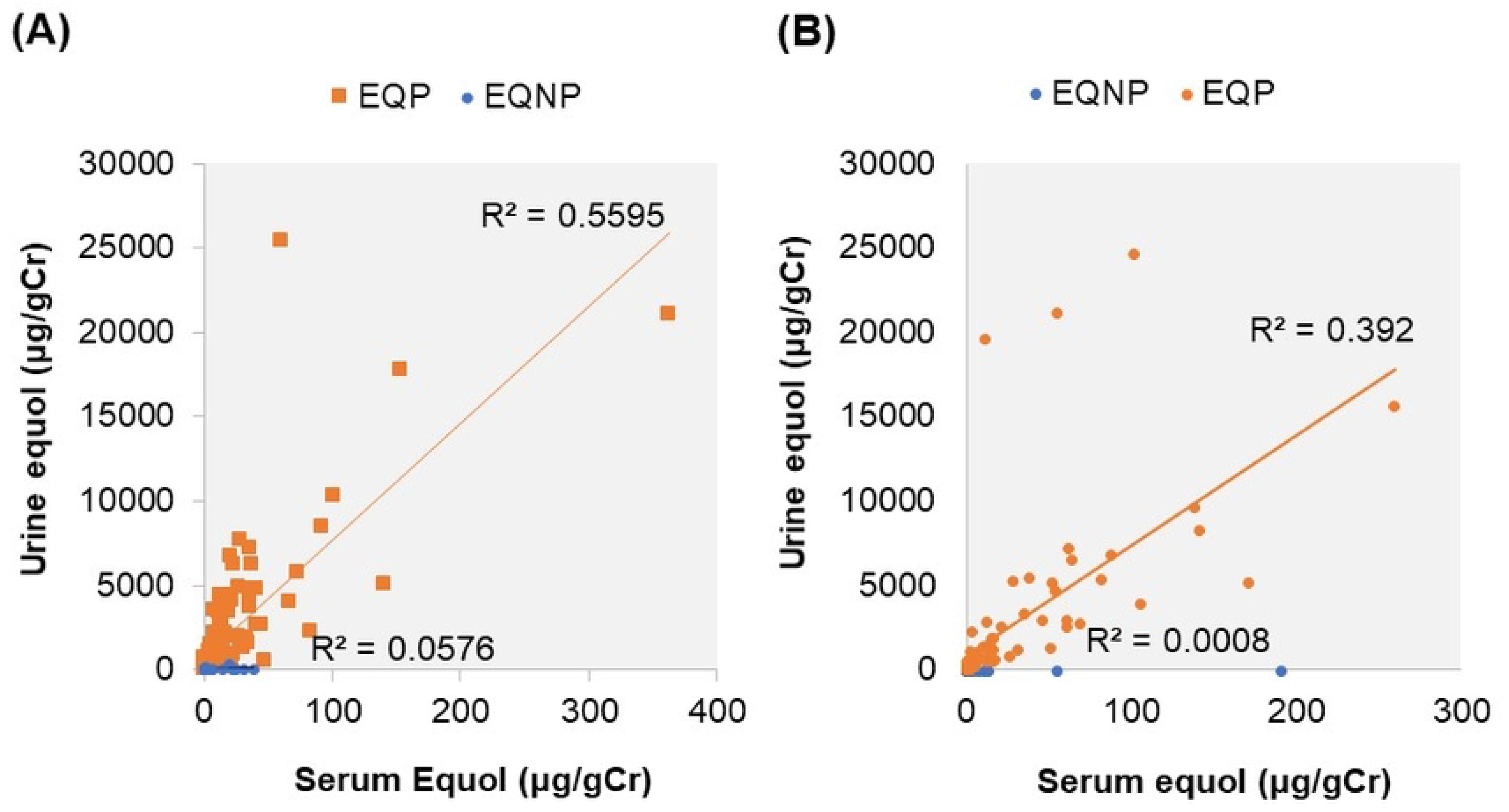
Relationship between serum and urine equol levels in male and female participants. Linear regression analysis in (A) male equol producers (n=114) and nonproducers (n=178), (B) female equol producers (n=81) and nonproducers (n=93). EQP=equol producers, EQNP=equol nonproducers.

### Comparison of other parameters between equol producers and nonproducers in men

**Table 2** shows that PSA levels in men were significantly lower in equol producers (0.8 v.s. 1.0 ng/ml, p=0.004), especially in male equol producers in their 40s (0.82 vs. 1.13 ng/ml, p<0.001) and 60 s (0.64 vs. 1.02 ng/ml, p<0.001), as shown in **Figure 3A**. In addition, a significant proportion of men with high LDL cholesterol levels were equol nonproducers (48.9% vs. 36.8%, p=0.043), and the proportions of equol producers were lower among those with high PSA levels (4.4% vs. 5.1%, p=0.068), as shown in **Figure 3B**.

**Fig 3.**
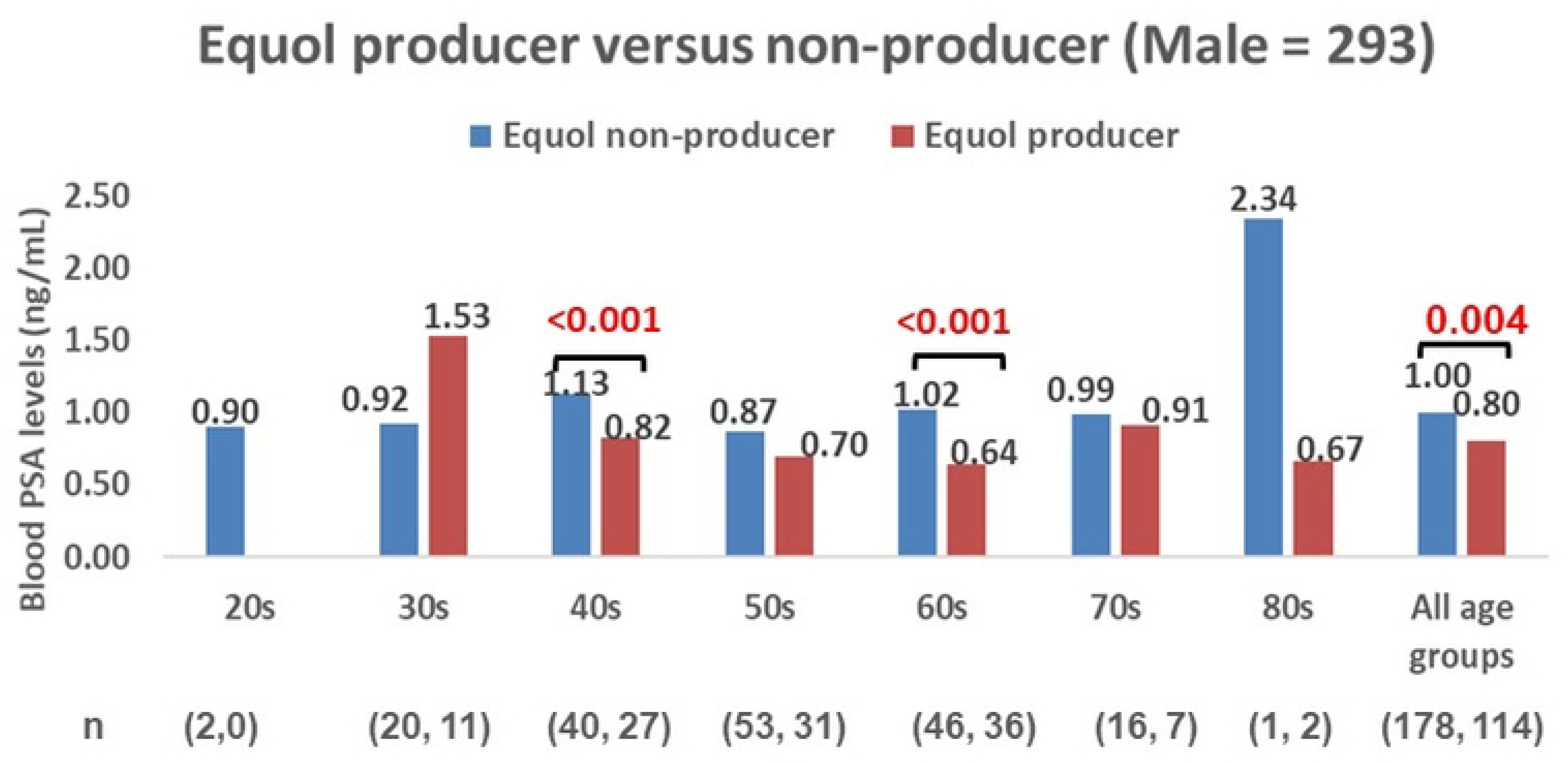
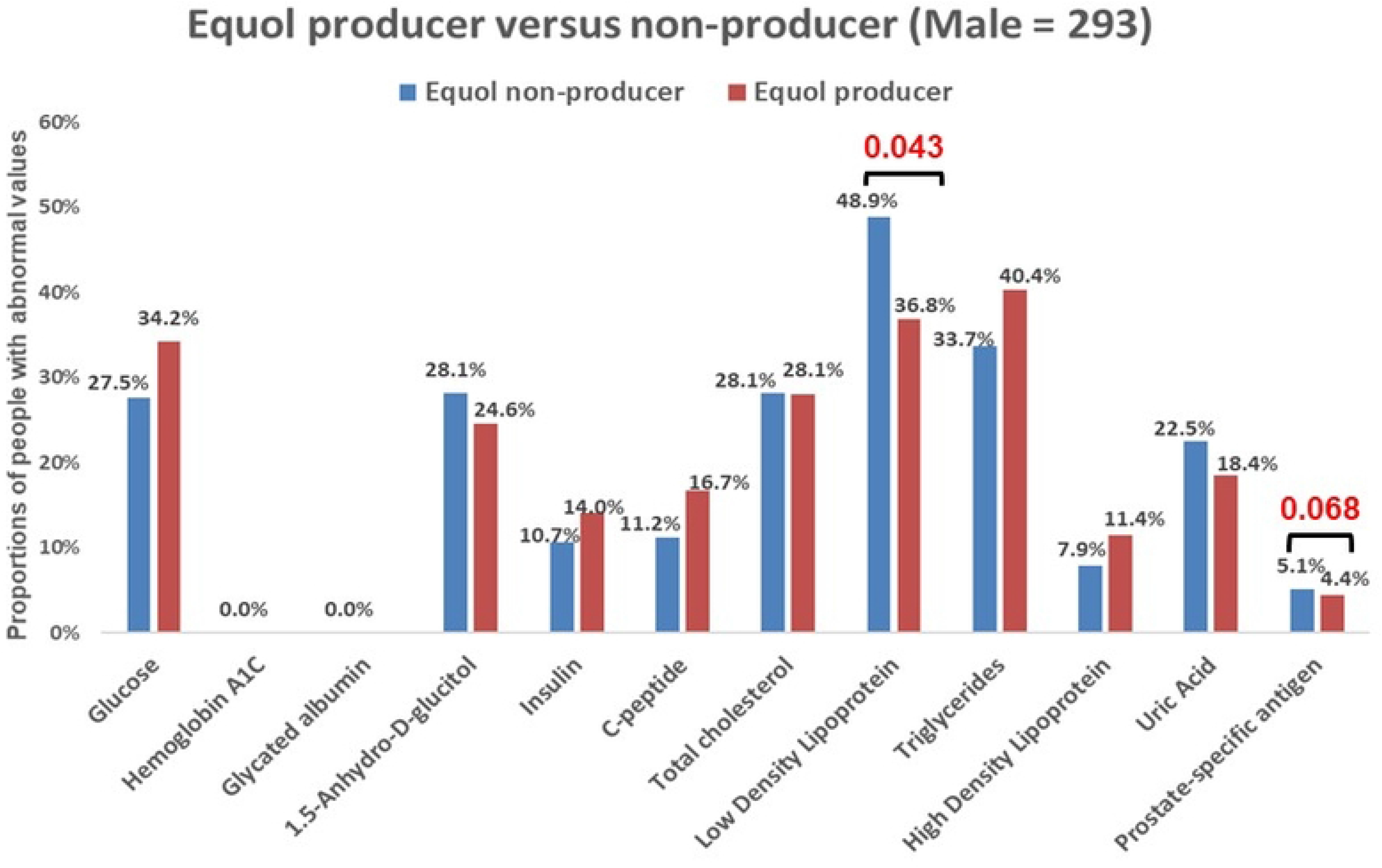
Comparison of blood parameters between equol producers and nonproducers in men. A: Comparisons of prostate-specific antigen levels between equol producers and nonproducers in each age group of men. PSA levels were expressed as medians and compared using the Mann‒Whitney U test. B: Comparisons of abnormal metabolic values between equol producers and nonproducers in men. The proportions of abnormal values were compared using the chi-squared test.

**Table 2:** Comparison between male equol producers and nonproducers.

|  | Equol producers (n=114) |  | Equol nonproducers (n=178) |  | p value |
| --- | --- | --- | --- | --- | --- |
| Age (years) |  |  |  |  | 0.6573<sup>a</sup> |
| 20s, n (%) | 0 | (0%) | 2 | (100%) |  |
| 30s, n (%) | 11 | (35%) | 20 | (65%) |  |
| 40s, n (%) | 27 | (40%) | 40 | (60%) |  |
| 50s, n (%) | 31 | (37%) | 53 | (63%) |  |
| 60s, n (%) | 36 | (44%) | 46 | (56%) |  |
| 70s, n (%) | 7 | (30%) | 16 | (70%) |  |
| 80s, n (%) | 2 | (67%) | 1 | (33%) |  |
| Serum equol (µg/gCr) | 7.64 | (0.64, 21.54) | 0.43 | (0.3, 0.72) | **<0.001** |
| Serum daidzein (µg/gCr) | 8.25 | (2.97, 28) | 13.01 | (4.85, 36.8365) | **0.025** |
| Urinary equol (µg/gCr) | 539.91 | (70.90, 2731.6) | 4.73 | (3.23, 8.23413) | **<0.001** |
| Urinary daidzein (µg/gCr) | 875.25 | (151.73, 2808.75) | 2152.50 | (956.15, 5324.83) | **<0.001** |
| log (Equol/Daidzein) | -0.30 | (-0.64, 0.2) | -2.57 | (-2.98, -2.0877) | **<0.001** |
| CRP | 0.05 | (0.05, 0.1) | 0.05 | (0.05, 0.1) | 0.752 |
| TSH | 1.51 | (1.06, 2.45) | 1.70 | (1.13, 2.5325) | 0.366 |
| FT4 | 1.31 | (1.20, 1.4) | 1.27 | (1.17, 1.4) | 0.559 |
| FT3 | 3.16 | (2.89, 3.4) | 3.13 | (2.88, 3.4175) | 0.907 |
| Glucose | 95.00 | (89.00, 103) | 94.00 | (88, 101.75) | 0.368 |
| HbA1C | 5.7% | (5.5%, 0.059) | 5.6% | (0.054, 0.059) | 0.074 |
| Glycated albumin | 0.13 | (0.13, 0.14) | 0.13 | (0.13, 0.14275) | 0.929 |
| 1.5-Anhydro-D-glucitol | 18.55 | (14.03, 22.9) | 18.85 | (13.13, 22.125) | 0.455 |
| Insulin | 5.20 | (3.73, 7.78) | 4.90 | (3.13, 7.3) | 0.472 |
| C-peptide | 1.65 | (1.23, 2.2) | 1.57 | (1.14, 2) | 0.215 |
| HOMA-IR | 1.23 | (0.85, 1.94) | 1.14 | (0.71, 1.81407) | 0.355 |
| Insulin resistance, n (%) | 18 | (16%) | 22 | (12%) | 0.511<sup>a</sup> |
| Total cholesterol | 192.50 | (173.00, 225.75) | 200.00 | (179.25, 226.5) | 0.291 |
| LDL | 109.00 | (96, 134) | 119.00 | (100.25, 140.75) | 0.114 |
| TG | 129.50 | (84.00, 187) | 107.50 | (72.25, 171.75) | 0.088 |
| HDL | 55.00 | (46.00, 63.75) | 56.00 | (47.25, 65.75) | 0.204 |
| UA | 6.20 | (5.43, 6.9) | 6.10 | (5.4, 6.9) | 0.899 |
| UCRE | 117.50 | (81.25, 174.5) | 125.00 | (80, 172.5) | 0.840 |
| PSA | 0.80 | (0.34, 1.3625) | 1.00 | (0.65, 1.53) | **0.004** |

Continuous variables are expressed as medians (interquartile ranges) and were compared by the Mann–Whitney U test except for <sup>a</sup>: chi-squared test; statistically significant p values are bold.

### Comparison between equol producers and nonproducers in women

Among women, no significant quantitative differences were observed between equol nonproducers except for isoflavone parameters (**Table 3**). However, equol nonproducers had more abnormal LDL cholesterol, triglyceride, and uric acid levels (**Figure 4**).

**Fig 4.**
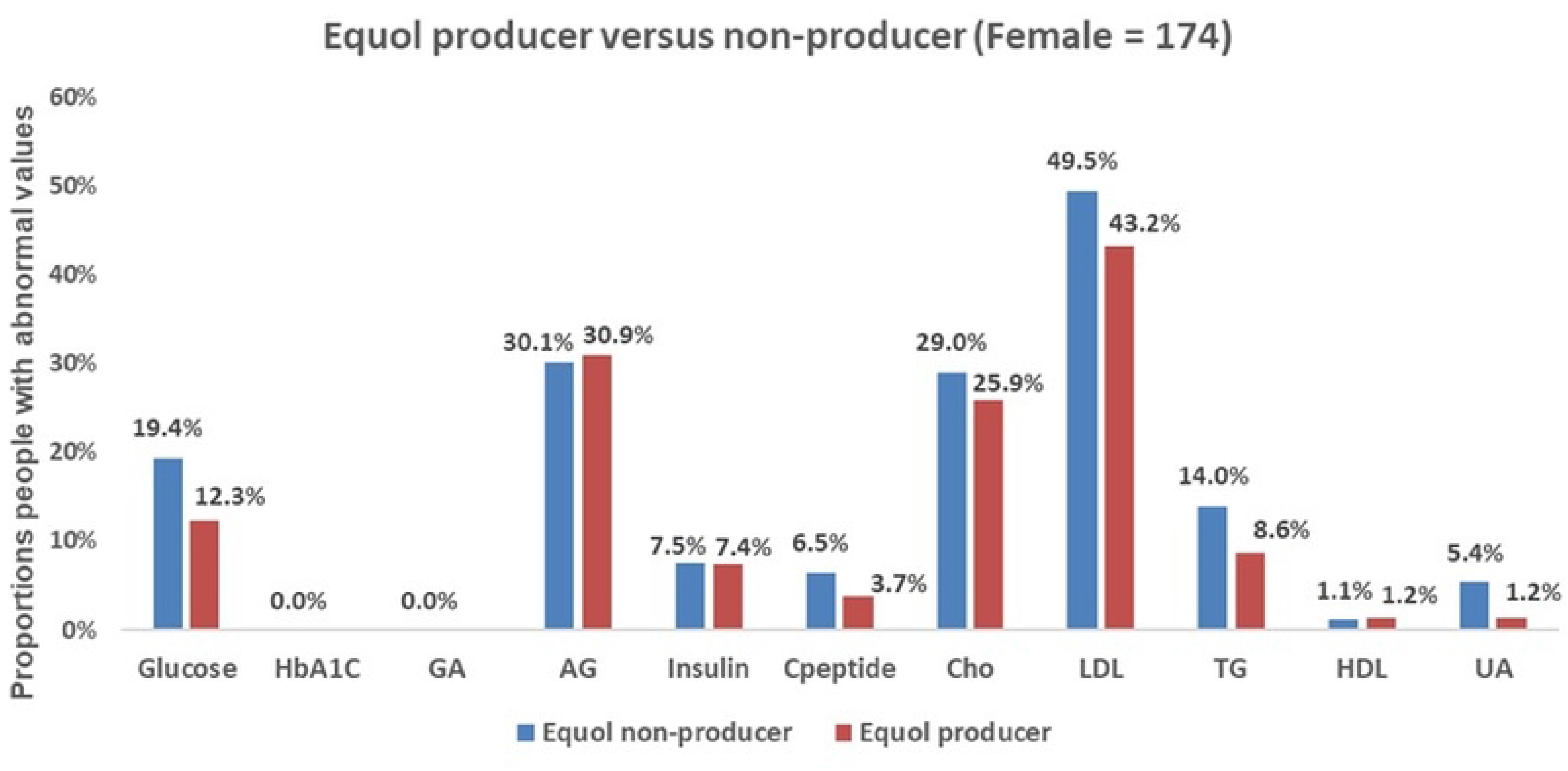
Comparison of blood parameters between equol producers and nonproducers in women. The proportions of abnormal values were compared using the chi-squared test.

**Table 3:** Comparison between female equol producers and nonproducers.

|  | Equol producers (n=81) |  | Equol nonproducers (n=93) |  | p value |
| --- | --- | --- | --- | --- | --- |
| Age |  |  |  |  | 0.8389<sup>a</sup> |
| 20s | 1 | (50%) | 1 | (50%) |  |
| 30s | 10 | (53%) | 9 | (47%) |  |
| 40s | 22 | (55%) | 18 | (45%) |  |
| 50s | 27 | (44%) | 34 | (56%) |  |
| 60s | 12 | (39%) | 19 | (61%) |  |
| 70s | 7 | (47%) | 8 | (53%) |  |
| 80s | 2 | (33%) | 4 | (67%) |  |
| Serum equol (µg/gCr) | 6.77 | (1.06, 30.93) | 0.71 | (0.52, 1.14) | **<0.001** |
| Serum daidzein (µg/gCr) | 13.67 | (5.73, 38.89) | 20.41 | (8.07, 51.81) | 0.1 |
| Urinary equol (µg/gCr) | 691.98 | (161.92, 2742.59) | 7.94 | (5.49, 12.2) | **<0.001** |
| Urinary daidzein (µg/gCr) | 1469.93 | (292.07, 3864.86) | 2907.89 | (1048.95, 7666.67) | **<0.01** |
| log (Equol/Daidzein) | -0.3 | (2.71, 3.54) | -2.46 | (2.79, 3.73) | **<0.001** |
| E2 | 0.25 | (0.25, 51.6) | 0.25 | (0.25, 70.5) | 0.74 |
| LH | 22.6 | (7.1, 35.2) | 22.55 | (6.6, 31.3) | 0.51 |
| FSH | 49.4 | (9.8, 76.2) | 47.65 | (6.3, 68.2) | 0.43 |
| CRP | 0.05 | (0.05, 0.1) | 0.05 | (0.05, 0.1) | 0.57 |
| TSH | 1.8 | (1.07, 2.79) | 1.83 | (1.33, 2.55) | 0.58 |
| FT4 | 1.22 | (1.14, 1.32) | 1.21 | (1.11, 1.31) | 0.3 |
| FT3 | 2.79 | (2.67, 3.09) | 2.8 | (2.59, 3.05) | 0.42 |
| Glucose | 90 | (86, 94) | 90 | (85, 97) | 0.64 |
| HbA1C | 0.056 | (0.054, 0.058) | 0.057 | (0.055, 0.059) | 0.05 |
| Glycated albumin | 0.138 | (0.131, 0.147) | 0.14 | (0.135, 0.148) | 0.17 |
| 1.5-Anhydro-D-glucitol | 15.9 | (13.5, 19.7) | 15.95 | (12.7, 19.4) | 0.55 |
| Insulin | 4.1 | (3, 4.9) | 4 | (2.5, 5.8) | 0.98 |
| C-peptide | 1.16 | (0.91, 1.58) | 1.155 | (0.92, 1.59) | 0.83 |
| HOMA-IR | 0.91 | (0.63, 1.16) | 0.91 | (0.55, 1.42) | 0.92 |
| Insulin resistance | 6 | (7.40%) | 8 | (9%) | 0.99<sup>a</sup> |
| Total cholesterol | 201 | (183, 220) | 202 | (178, 224) | 0.86 |
| LDL | 115 | (98, 137) | 116.5 | (96, 137) | 0.61 |
| TG | 70 | (55, 105) | 69.5 | (51, 110) | 0.84 |
| HDL | 71 | (58, 80) | 69 | (55, 76) | 0.3 |
| UA | 4.6 | (4, 5.4) | 4.65 | (3.9, 5.3) | 0.9 |
| UCRE | 71 | (51, 116) | 72 | (50, 100) | 0.63 |

Continuous variables are expressed as medians (interquartile ranges) and were compared by the Mann–Whitney U test except for <sup>a</sup>: chi-squared test; statistically significant p values are bold.

## Discussion

We found that the cut-off value of the log-transformed urinary equol to daidzein ratio was almost consistent in both sexes. Previously, this cut-off value was reported in female participants only [28]. Therefore, this is the first study that could reproduce the same results in both men and women. Additionally, we found that urinary and serum equol concentrations were significantly correlated in equol producers but not in nonproducers. This also highlighted the important concept that it could be difficult to differentiate the equol producer phenotypes relying on either absolute serum or urinary equol concentrations. Therefore, Setchell et al proposed a robust method of defining an equol producer based upon the precursor-product relation using the log10-transformed ratio of urinary equol to its precursor daidzein [29].

The cut-off value of equol producers in Setchell’s study was -1.75 after a standard 3-day challenge of soy foods containing isoflavones. Here, in our study, we used -1.42 as the cut-off value, as we did not perform the soy challenge, to reflect the real-time measurement as in the Japan Nurses’ Health Study [28]. By using both the precursor and product relation to determine the equol producer phenotype, we also minimized errors due to large variance in equol concentrations due to differences in dietary isoflavone intake, pharmacokinetics, and methodologies for measuring the intrinsic equol. This definition identified 195 participants among 466 as equol producers (42%), which was similar to the Japan Nurses’ Health Study on 4,412 participants that has used the same cut-off value (41.5%). The proportion of equol producers in females was larger than that in males (47% versus 39%). The lenient nature of the relationship between serum and urinary equol levels in women might have reflected the stable serum equol concentration in women.

Studies on the benefits of soy isoflavones have yielded inconsistent results. This could be most likely due to the variations in equol producer phenotypes. Even in the studies that assessed the equol producer phenotypes, some results failed to reach statistical significance due to small sample sizes. For example, in this study, female equol nonproducers tended to have higher LDL cholesterol, triglyceride, high sensitivity C-reactive protein and uric acid levels, but the results were not statistically significant. However, in our previous study on 743 healthy women, equol producers in their 50s and 60 s, the age groups with declining estrogen levels, had favorable blood levels of lipids, uric acid, bone resorption markers, high sensitivity C-reactive protein, and homocysteine [30]. These positive effects were due to the estrogenic and antioxidant action of equol.

In this study, male equol producers had significantly lower PSA levels than nonproducers, especially in men in their 40s and 60 s. Additionally, abnormal PSA values were rarely associated with male equol producers. Several studies have reported that equol can decrease serum PSA levels by its antiandrogenic action on 5-alpha-dehydro testosterone, decrease prostate size, and thus reduce the risk of prostate cancer [12, 13, 31,32]. Therefore, this study has also added knowledge to equol benefits on men’s prostate health. Additionally, it was a significant finding that abnormal LDL cholesterol levels were associated with male equol nonproducers. Furthermore, male equol nonproducers tended to have abnormal HDL cholesterol and uric acid levels, which needs to be explored in larger studies.

This study has the following limitations. First, we did not have the detailed characteristics of the study participants, including medical history, anthropometric measures, and dietary habits. However, it would not affect the results of this epidemiological study significantly, although they might provide more information. Second, the sample size varied among the different age groups. Age is one of the most important determinants affecting the parameters that we assessed. Therefore, we need to be cautious about generalizing the results in all age groups and need to consider further studies based on the sample size calculations using the effect sizes in this study.

## Conclusion

This is the first study to confirm the use of the precursor-product relation as a robust cut-off value to identify the equol producer phenotype in box men and women using the LC‒MS/MS method. It is also the first study that examined the differences between urine and serum equol concentrations in both females and males in equol producers and nonproducers. The equol-producing ability tended to be higher in females, suggesting a relationship between estrogen and equol-producing ability. In males, equol-producing subjects had significantly lower PSA levels, suggesting a relationship between equol-producing ability and reduced risk of prostate disease. In addition, both female and male equol producers have positive effects on blood lipids and uric acid levels. However, we need more robust clinical trials to examine the health benefits of equol in both men and women.

## Data Availability

Data cannot be shared publicly because of the restriction from the Kyoto University Hospital. Data are available only after negotiating with the Kyoto University to allow access after submitting the proposal to the Institutional Review Board of the University the Kyoto University has designated for researchers who meet the criteria for access to confidential data.

## Acknowledgements

We would like to acknowledge all the participants who have consented to use their health screening data for the purpose of medical research, Himedic Kyoto University Hospital, and all the persons who have contributed to making this research possible.

## Authors’ contributions

RY is the principal investigator who conceptualized or designed the entire study, conducted data analysis and drafted the manuscript.

KZYM (ORCID: 0000-0001-5553-1245) was the corresponding author, involved in data analysis and interpretation and revised the manuscript.

JT is the coinvestigator who contributed to the conceptualization and implementation of the study and revision of the manuscript.

## Notes

### Competing Interest Statement

The authors have declared no competing interest.

### Funding Statement

NO. The funders had no role in study design, data collection and analysis, decision to publish, or preparation of the manuscript.

### Author Declarations

The study was approved by the Institutional Review Board of The University of Tokyo

